# Improving Automated Deep Phenotyping Through Large Language Models Using Retrieval Augmented Generation

**DOI:** 10.1101/2024.12.01.24318253

**Authors:** Brandon T. Garcia, Lauren Westerfield, Priya Yelemali, Nikhita Gogate, E. Andres Rivera-Munoz, Haowei Du, Moez Dawood, Angad Jolly, James R. Lupski, Jennifer E. Posey

**Affiliations:** Department of Molecular and Human Genetics, Baylor College of Medicine, Houston, TX, 77030, USA; Medical Scientist Training Program, Baylor College of Medicine, Houston, TX, 77030, USA; Genetics and Genomics Graduate Program, Baylor College of Medicine. Houston, TX, 77030, USA; Texas Children’s Hospital, Houston, TX, 77303, USA; Human Genome Sequencing Center, Baylor College of Medicine, Houston, TX, 77030, USA; Department of Pediatrics, Baylor College of Medicine, Houston, TX, 77030, USA

**Keywords:** Large language models (LLMs), Retrieval augmented generation (RAG), Phenotyping, Human Phenotype Ontology (HPO), Natural Language Processing (NLP), Clinical Genomics, Generative Pre-trained Transformer (GPT), Generative AI, Llama-3

## Abstract

**Background:** Diagnosing rare genetic disorders relies on precise phenotypic and genotypic analysis, with the Human Phenotype Ontology (HPO) providing a standardized language for capturing clinical phenotypes. Traditional HPO tools, such as Doc2HPO and ClinPhen, employ concept recognition to automate phenotype extraction but struggle with incomplete phenotype assignment, often requiring intensive manual review. While large language models (LLMs) hold promise for more context-driven phenotype extraction, they are prone to errors and “hallucinations,” making them less reliable without further refinement. We present RAG-HPO, a Python-based tool that leverages Retrieval-Augmented Generation (RAG) to elevate LLM accuracy in HPO term assignment, bypassing the limitations of baseline models while avoiding the time and resource intensive process of fine-tuning. RAG-HPO integrates a dynamic vector database, allowing real-time retrieval and contextual matching.

**Methods:** The high-dimensional vector database utilized by RAG-HPO includes >54,000 phenotypic phrases mapped to HPO IDs, derived from the HPO database and supplemented with additional validated phrases. The RAG-HPO workflow uses an LLM to first extract phenotypic phrases that are then matched via semantic similarity to entries within a vector database before providing best term matches back to the LLM as context for final HPO term assignment. A benchmarking dataset of 120 published case reports with 1,792 manually-assigned HPO terms was developed, and the performance of RAG-HPO measured against existing published tools Doc2HPO, ClinPhen, and FastHPOCR.

**Results:** In evaluations, RAG-HPO, powered by Llama-3 70B and applied to a set of 120 case reports, achieved a mean precision of 0.84, recall of 0.78, and an F1 score of 0.80—significantly surpassing conventional tools (p<0.00001). False positive HPO term identification occurred for 15.8% (256/1,624) of terms, of which only 2.7% (7/256) represented hallucinations, and 33.6% (86/256) unrelated terms; the remainder of false positives (63.7%, 163/256) were relative terms of the target term.

**Conclusions:** RAG-HPO is a user-friendly, adaptable tool designed for secure evaluation of clinical text and outperforms standard HPO-matching tools in precision, recall, and F1. Its enhanced precision and recall represent a substantial advancement in phenotypic analysis, accelerating the identification of genetic mechanisms underlying rare diseases and driving progress in genetic research and clinical genomics.

## Introduction

In genomic medicine and research, phenotypic and genotypic analyses are critical for achieving accurate molecular diagnoses. Deep phenotyping allows for a detailed understanding of a patient’s clinical presentation, which can then be matched to potential genetic causes. [1] Genomic analysis provides the molecular insights necessary to identify pathogenic variants that may be contributing to disease. [2] Together, these approaches facilitate a comprehensive evaluation of patients, particularly those with rare or undiagnosed conditions, and offer the possibility of uncovering genetic etiologies that might otherwise remain elusive. [3] Integrating clinical and molecular data is essential for providing patients with concrete answers, guiding their treatment, and improving outcomes. [4–7]

The Human Phenotype Ontology (HPO) is a standardized vocabulary with a hierarchical structure essential for deep phenotypic analysis. [8, 9] The HPO’s hierarchical structure allows clinicians and researchers to consistently compare and categorize phenotypes by connecting general phenotypic terms with more specific terms, enabling the capture of subtle differences across individuals despite observed variability in patient presentations and the nomenclature used to describe them. [10] The ID numbers associated with specific HPO terms support computational comparisons of phenotypically similar individuals within large cohorts, enabling more precise matching of phenotypes to genetic variants. [8, 11, 12] With over 17,000 terms, HPO enhances the capacity for machine-readable data, paving the way for the advanced phenotypic matching tools necessary in genomic medicine today.

While HPO terms provide a standardized framework for cataloging and investigating patient phenotypes, the process of deep phenotyping remains labor-intensive and reliant on clinical expertise. Many tools, such as Doc2HPO, ClinPhen, and FastHPOCR, seek to automate the extraction of relevant phenotypic phrases from clinical records using concept recognition methods. [11–13] These tools employ dictionary-based approaches to match clinical text with HPO terms, facilitating the identification of key phenotypes. However, despite their utility, these tools often miss substantial portions of patient phenotypes, necessitating thorough manual cross-examination to ensure accurate phenotype annotation.

Large language models (LLMs) are advanced computational programs trained to understand and generate natural language based on patterns present in human-generated text. As part of the increased popularity of generative Artificial Intelligence (AI), these programs offer a promising opportunity to significantly improve automated deep phenotyping. [14, 15] Within the biomedical field, remarkable advances in LLM technology have led to rapid growth in the use of technology in various clinical and basic science applications, including interpretation of radiological results or analyzing medical text. [16–20] The ability of LLMs to understand natural language context is a critical component in improving our ability to extract relevant clinical phenotypes from patient data. Consequently, LLMs have entered the phenotypic analysis space through tools like PhenoTagger, PhenoBERT, and PhenoGPT. [21, 22]

However, the integration of LLMs into phenotypic analysis presents new challenges. As other authors have noted, currently available LLMs are resource-intensive and slow in their reasoning compared to other machine learning methods. [13] Importantly, they are also prone to hallucinations, in which the model generates incorrect information and confidently asserts it as fact. [23] Many LLM-based tools for phenotypic analysis address these concerns through fine-tuning, which involves further training of a model with additional data. [21] While this process increases the accuracy of LLM responses, it is computationally expensive (requiring substantial GPU and RAM), time-consuming, and necessitates specialized expertise in working with generative AI models. [14, 23, 24] With the rapid advancement of LLM technology and continual updates to the HPO lexicon, fine-tuning LLMs for phenotypic analysis becomes impractical for most clinicians and researchers involved in diagnosing rare genetic diseases.

Alternatively, retrieval-augmented generation (RAG) is a practical solution to the limitations of fine-tuning. RAG uses vector databases to retrieve relevant information from source documents in real-time, making it easier and faster to update the system with new information. [16, 23, 25, 26] Users can refresh the underlying vector database with minimal effort, allowing the system to stay current without the need to retrain the LLM. This makes RAG more cost-effective and adaptable, particularly for users without advanced technical expertise and who use a resource that constantly updates, while still leveraging the power of LLMs to enhance the accuracy and precision of results.

We have developed a Python-based program, RAG-HPO, to apply RAG to deep phenotyping, drastically increasing LLMs’ ability to accurately assign HPO terms to patient phenotypes. This enables us to leverage the LLMs’ superior ability to extract phenotypic information from clinical data, improving the process of automated deep phenotyping. RAG-HPO is simple to use, does not require extensive understanding of LLMs, works with any language model, and does not require computationally intense resources like GPUs and large amounts of RAM for use. By utilizing a vector database, RAG-HPO can be easily updated with new information from the HPO database and user input. When paired with Llama-3, RAG-HPO demonstrates superior precision and recall compared to popular dictionary-based concept recognition tools.

## Methods

RAG-HPO is a Python-based tool designed to extract clinical phenotypes from medical free-text and assign HPO terms to those phrases using RAG. Users can employ any LLM of their choosing by providing an Application Programming Interface (API) key. Below, we describe implementation of this tool with the LLM Llama-3.1 70B for the benchmarking of RAG-HPO.

### Data Preparation, Embedding, and Vectorization

The vector database used by RAG-HPO utilizes a python dictionary with key-value pairs, where keys are clinical words and phrases that reliably match to HPO ID numerical values (e.g. furrowed tongue: HP:0000221). The initial dictionary was extracted from the HPO database and contains HPO term titles, names, definitions, and synonyms paired with the HPO ID. The initial dictionary was supplemented with additional values that reliably mapped to specific HPO IDs and validated using orthogonal approaches, including established HPO analysis tools and manual annotations. Currently, this custom dictionary contains over 54,000 unique value phrases that correspond to individual HPO IDs stored in JavaScript Object Notation (JSON) format [See **Additional File 1**]. The refined dataset serves as the basis for the vector database used in RAG-HPO. Fast Embed was employed to convert each term within the JSON file to a high-dimensional vector to capture semantic relationships between phenotypic terms and factor together lineage information to bring like terms closer to one another, enabling enhanced similarity searches during phenotypic matching (**Figure 1**).

**Figure 1:**
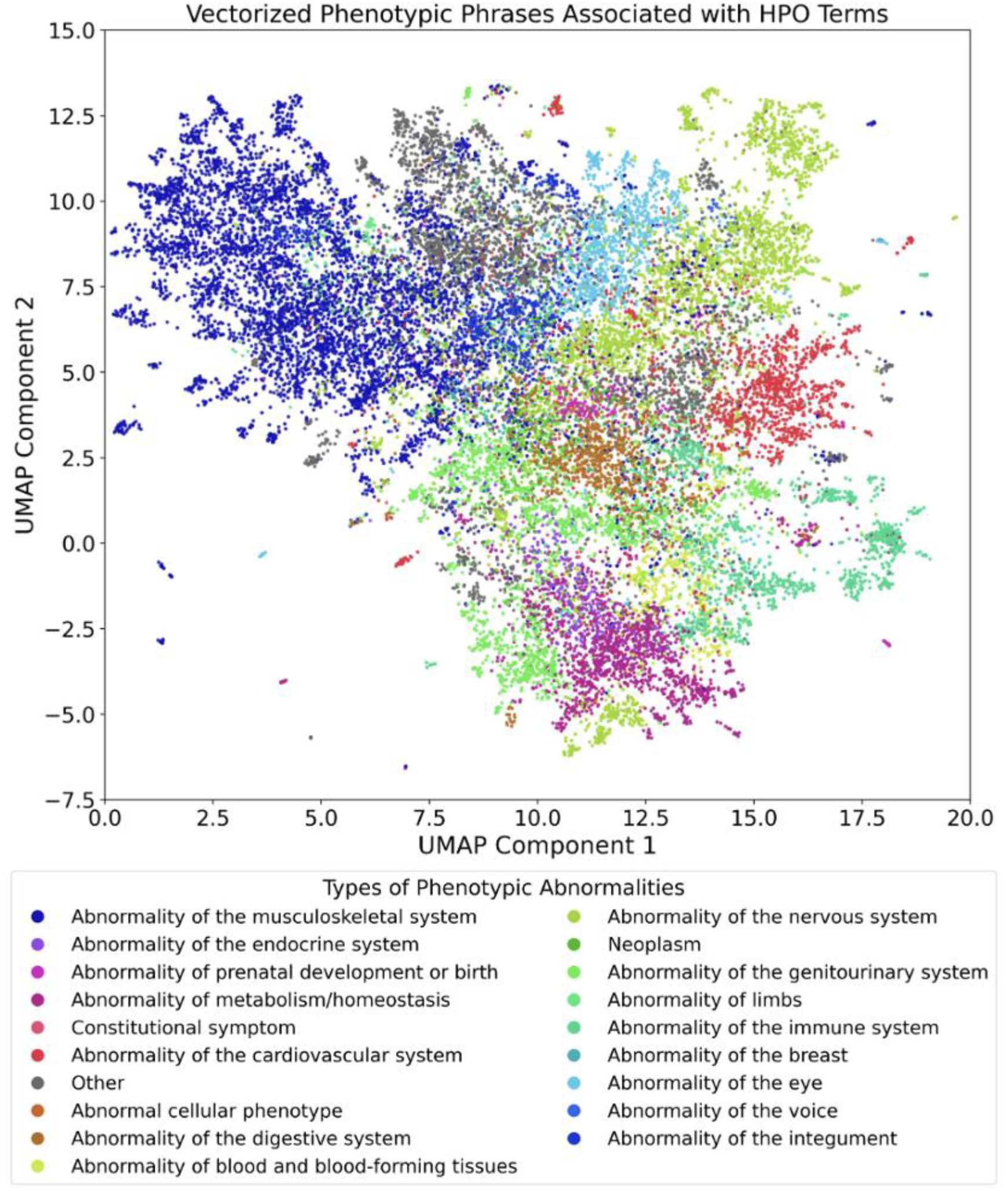
*UMAP representation of vector database storage of the Human Phenotype Ontology.* The HPO database was downloaded, and key information was extracted and restructured for vectorization, including the HPO ID, Title, Definition, Comments, and Synonyms. Additional phrases were created using LLMs and validated using current HPO analysis tools to increase the robustness of the database. The current database contains 54,000 phrases that map to specific HPO IDs.

The embedded database is stored as a NumPy array for use by RAG-HPO, serving as a resource to support the LLM in making accurate assignments. When the analysis program is initiated, the NumPy array is indexed in a vector database optimized for dense vector retrieval, allowing for efficient approximate nearest-neighbor searches. Relevant metadata is also indexed within the database to assist the LLM in reasoning through the assignment process.

### RAG-HPO Workflow

The program processes free-text provided as strings, converting them into HPO terms. Input text can either be supplied directly by the user or batch-processed from a CSV file containing clinical notes. The text is first passed to the user’s chosen LLM via an API call, where a custom system prompt instructs the LLM to extract phenotypic phrases that describe the patient’s health status, while disregarding non-relevant information such as administrative details or general observations that do not pertain to the patient’s health (**Figure 2A, Additional File 2**).

**Figure 2:**
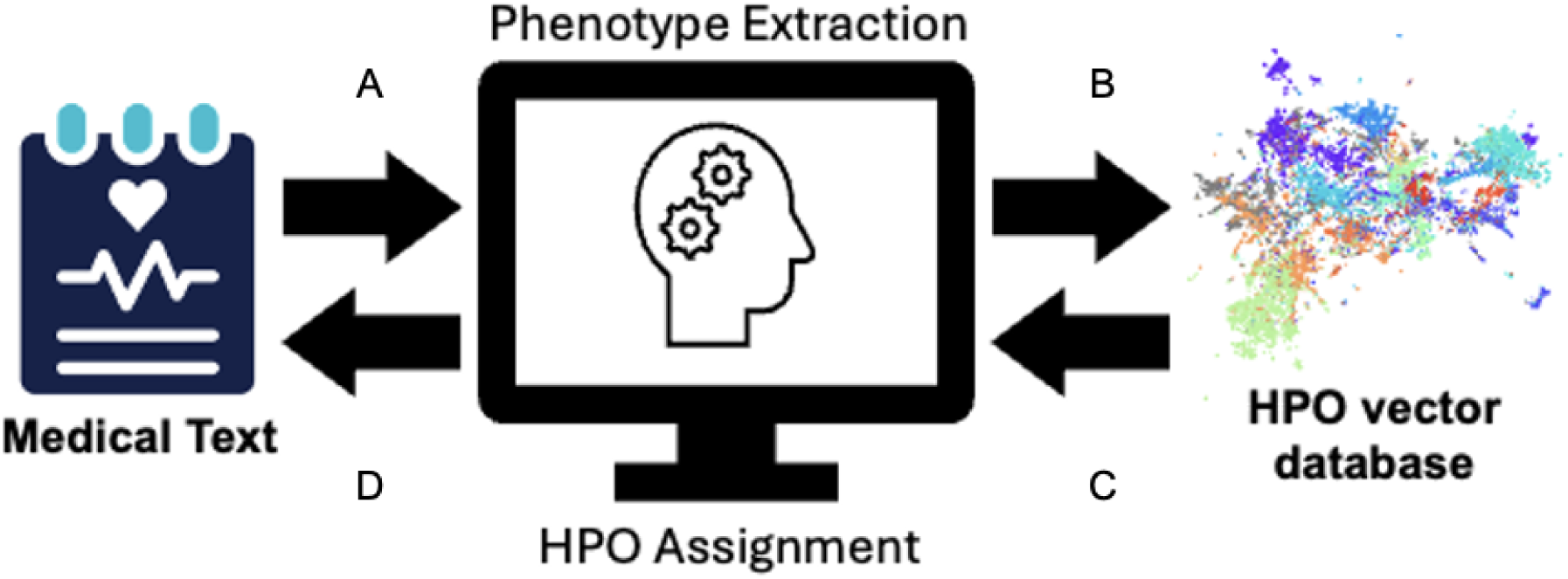
*RAG-HPO extracts phenotypic information and returns HPO terms*. RAG-HPO works in two phases, phenotype extraction and HPO assignment, to determine the appropriate HPO terms for the evaluated free clinical text. In the first phase, the clinical information is parsed to the LLM for extraction of clinical abnormalities (A). The extracted phrases are then vectorized and compared to the HPO vector database using semantic similarity search (B). In the second phase, the top 20 most similar phrases for each original extracted phrase are then returned to the LLM for assignment of HPO terms (C). Once all extracted phrases are analyzed, the list of HPO terms is returned to the user for verification and downstream analysis (D).

Once extracted, these phenotypic phrases are prepared for the RAG process (**Figure 2B**). To optimize computational efficiency and reduce the workload on the LLM, each extracted phrase is first compared to the metadata within the embeddings array using fuzzy matching. Exact matches are directly included in the results without further processing, while phrases that do not have an exact match are converted into high-dimensional semantic vectors using Fast Embed. The generated embeddings, along with associated metadata (such as HPO terms, lineage information, and organ system), are compared to the developed HPO database in a similarity search via Facebook AI Similarity Search, an efficient package for indexing and searching vector data, to identify related HPO terms (**Figure 2B**). The program retrieves the top 20 most semantically similar vectors, along with their associated metadata, including the relevant HPO terms, lineage, and other contextual information. Then, the surrounding sentence from the original clinical text is retrieved to provide additional context for understanding the extracted phrase.

The extracted clinical phrases are resubmitted to the LLM, which uses the additional context provided by the metadata to select the most appropriate and detailed HPO term representing the most distal matching node (highest possible information content [27]) in the ontology (**Figure 2C**). After each extracted phrase and metadata block are passed through the LLM, a completed list of HPO terms for the whole passage is returned to the user (**Figure 2D**). When analyzing batches, the results are saved as a JSON object list within a copy of the original CSV file under a new column.

### Performance Evaluation of RAG-HPO and Other HPO Analysis Tools

RAG-HPO, ClinPhen, Doc2HPO, and FastHPOCR were evaluated based on their ability to accurately extract HPO terms from previously published case reports. The test cohort used was derived from 120 case-reports sourced from reputable medical journals. Case selection was carried out by a clinician independent of the evaluation process to mitigate selection bias and ensure the inclusion of a broad range of medical specialties. Manual annotation of HPO terms was conducted by a certified genetic counselor and clinically trained graduate/medical student. Cases with fewer than three manually identified HPO terms were excluded, resulting in a final cohort of 112 cases containing a total of 1,792 HPO terms and a mean of 16 terms per case. The average length of each case report was 400 words. The manually annotated standard was used as the baseline for comparison across multiple tools, including ClinPhen, Doc2HPO, and FastHPOCR. For RAG-HPO, the LLAMA-3 70B model was employed as the underlying LLM. Additionally, to assess the specific contribution of RAG-HPO to the performance, the base performance of LLAMA-3 70B on a subset of cases is included, without the RAG process, to highlight the impact of RAG-HPO on improving precision and recall in HPO term assignment by an LLM.

In our analysis, we considered any HPO term called by an analysis tool to be a true positive if it matched the manually annotated standard. False positives were then any called HPO term that did not match manually annotated terms. Failure to identify an HPO term from the standard was considered a false negative. The number of true positives, false positives, and false negatives were used to calculate three key metrics: precision, recall, and the F1 score. These scores were used in determining the performance of the HPO analysis tools.

Precision measures the accuracy of each tool in returning relevant HPO terms, and is calculated as:

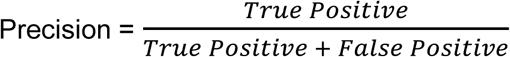

Recall measures the ability of the tool to retrieve all HPO terms present within a case note, and is calculated as:

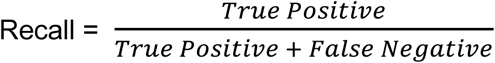

The F1 score is a weighted average of precision and recall on a scale of 0-1 and was calculated using the following formula:

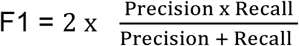

## Results

### Retrieval-Augmented Generation greatly increases LLM’s ability to identify correct HPO terms

The goal of RAG is to enhance a language model’s ability to retrieve and generate relevant information, thereby reducing the occurrence of “hallucinations”—a common issue where LLMs produce false or inaccurate information. In the context of HPO analysis, hallucinations typically manifest as incorrect or non-existent HPO terms that fail to correspond accurately to the clinical phrases being analyzed.

To evaluate the effectiveness of RAG-HPO in improving LLM performance in HPO analysis, we analyzed a cohort of 77 cases and compared the performance of Llama-3 70B both with and without RAG integration (**Table 1, Additional File 3**). When using the Llama-3 70B model alone, precision, recall, and F1 scores were 0.13, 0.12, and 0.12, respectively. However, when combined with RAG-HPO, these metrics improved dramatically, yielding a precision of 0.84, recall of 0.78, and an F1 score of 0.80. These results confirm that RAG significantly enhances LLM performance in HPO analysis, with the RAG component directly responsible for the improvements in accuracy (**Figure 3A**).

**Figure 3:**
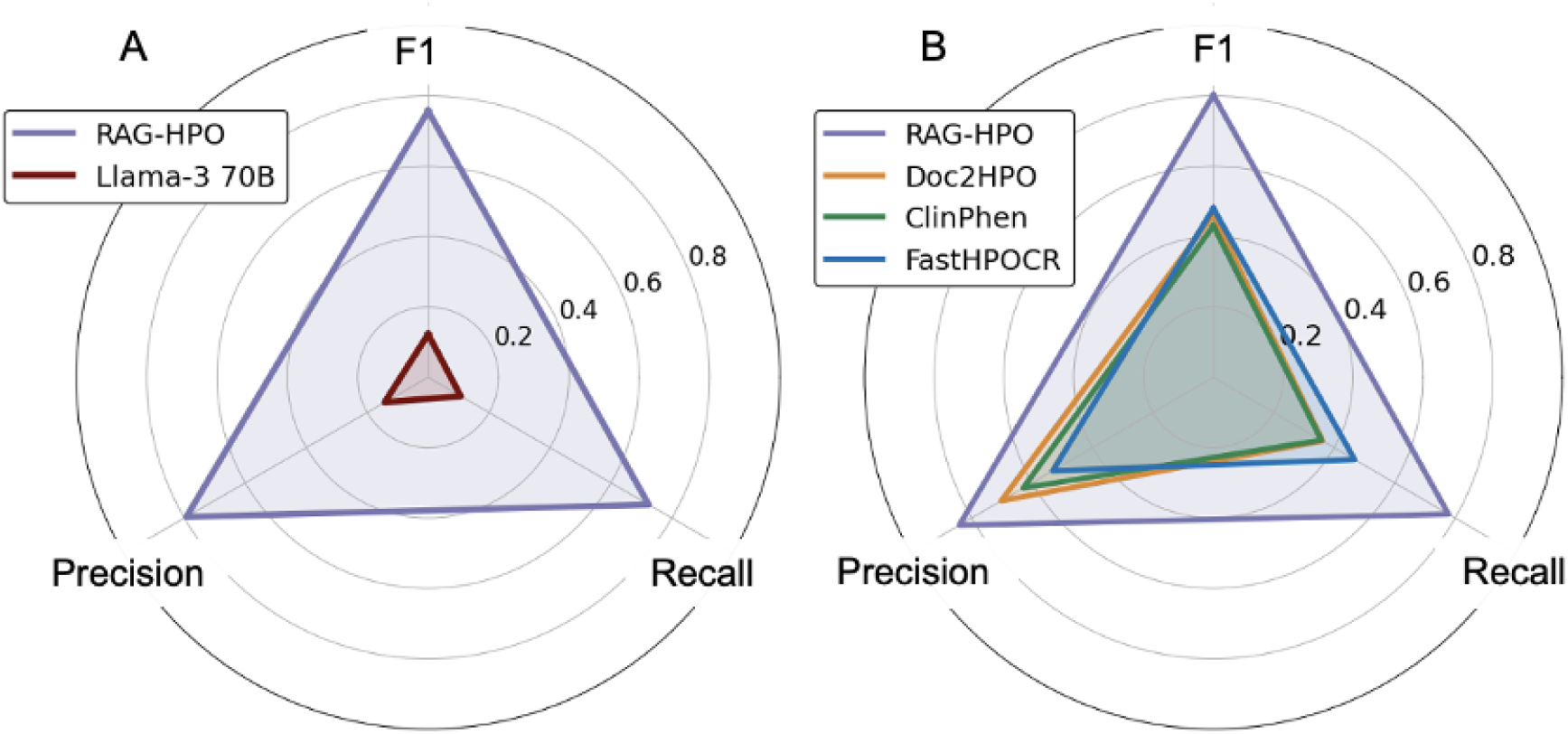
*RAG-HPO has superior recall and precision compared to established HPO analysis tools.* A) We compared the output of Llama-3 70B alone to RAG-HPO paired with Llama-3 70B in assigning HPO terms to 20 previously published case reports. Retrieval Augmented Generation greatly improved the ability of large language models to accurately assign HPO terms (F1 of .12 vs 78 respectively, *p< .0001)*. B) We then compared the capabilities of RAG-HPO paired with Llama3-70B to Doc2HPO, ClinPhen, and FastHPOCR on a cohort of 112 previously published cases. As illustrated by these scores, RAG-HPO demonstrates significantly better performance in precision, recall, and F1 score compared to other analysis tools.

**Table 1.** Retrieval augmented generation improves large language models’ ability to accurately assign HPO terms.

| Analysis Tool | True Pos | False Pos | False Neg | Precision | Recall | F1 | T test Comparison |  |  |
| --- | --- | --- | --- | --- | --- | --- | --- | --- | --- |
|  |  |  |  |  |  |  | Precision | Recall | F1 |
| **RAG-HPO** | 223 | 58 | 85 | 0.84 | 0.78 | 0.80 | *** | *** | *** |
| **Llama-3 70B** | 33 | 198 | 275 | 0.13 | 0.12 | 0.12 |  |  |  |

In a cohort of 77 cases, RAG-HPO with Llama-3 70B correctly identified 223 out of 308 available HPO terms. Llama-3 70B alone only identified 33 available terms. While Llama-3 70B was able to correctly identify phenotypic phrases at the same rate of RAG-HPO, its HPO term assignments were fraught with hallucinations and unrelated HPO terms. \*\*\* indicates $p$ value < .0001

### RAG-HPO Versus Established HPO Analysis Tools

Using the previously described annotated cohort, RAG-HPO’s performance was compared to established HPO extraction tools, specifically ClinPhen, Doc2HPO, and FastHPOCR. RAG-HPO leverages the LLAMA-3 70B model with RAG to enhance precision and recall in HPO term identification (**Table 2**).

**Table 2.** RAG-HPO enables LLMs to have greater precision and recall compared to traditional dictionary-based concept recognition approaches.

| Analysis Tool | True Pos | False Pos | False Neg | Precision | Recall | F1 | Comparison to RAG-HPO |  |  |
| --- | --- | --- | --- | --- | --- | --- | --- | --- | --- |
|  |  |  |  |  |  |  | Precision | Recall | F1 |
| **RAG-HPO** | 1368 | 257 | 424 | 0.84 | 0.78 | 0.80 | N/A | N/A | N/A |
| **Doc2HPO** | 643 | 251 | 1150 | 0.70 | 0.36 | 0.46 | **** | **** | **** |
| **ClinPhen** | 642 | 364 | 1150 | 0.63 | 0.36 | 0.43 | **** | **** | **** |
| **FastHPOCR** | 820 | 721 | 973 | 0.53 | 0.47 | 0.48 | **** | **** | **** |
| RAG-HPO paired with Llama-3 70B correctly identified significantly more HPO terms compared to other assessed programs with a smaller effective false positive rate. **** indicates $p < 0.00001$. Abbreviations: Neg, negative; Pos, positive. | | | | | | | | | |

For this cohort, RAG-HPO achieved significantly higher precision, recall, and F1 scores compared to the other HPO analysis tools (**Figure 3B**) with p<0.00001 (**Table 2, Table S1 of Additional File 3**). The most notable strength of RAG-HPO was its ability to identify twice as many correct HPO terms from the test cases as any other method tested. On average, RAG-HPO correctly identified a mean of 12 HPO terms per case, while Doc2HPO and ClinPhen averaged 6 HPO terms each. FastHPOCR identified a mean of 7 HPO terms per case but had a significantly higher false positive rate, resulting in a modest increase in recall at the expense of lower precision.

RAG-HPO also achieved the highest precision score of the group (0.84). Doc2HPO and ClinPhen yielded precision scores of 0.70 and 0.63, respectively, which aligned with their performance as described in the literature. While FastHPOCR identified nearly as many HPO terms as RAG-HPO, its precision was notably lower (0.53), meaning that only a little over half of the terms identified by FastHPOCR were exact matches to the manually curated standard.

As an example, we have included a small representative case to illustrate the strengths and weaknesses of the analyzed programs. **Figure 4** shows the phenotypic phrases identified by each HPO analysis program, while **Table 3** provides a detailed breakdown of the HPO terms assigned to these phrases by each program. In this case, RAG-HPO identified 8 phenotypic phrases and correctly assigned 7 HPO terms. Doc2HPO identified 4 phrases, 2 of which were correct; ClinPhen identified 2 phrases, both of which were correct; and FastHPOCR identified 10 phrases, 4 of which were correct.

**Figure 4:**
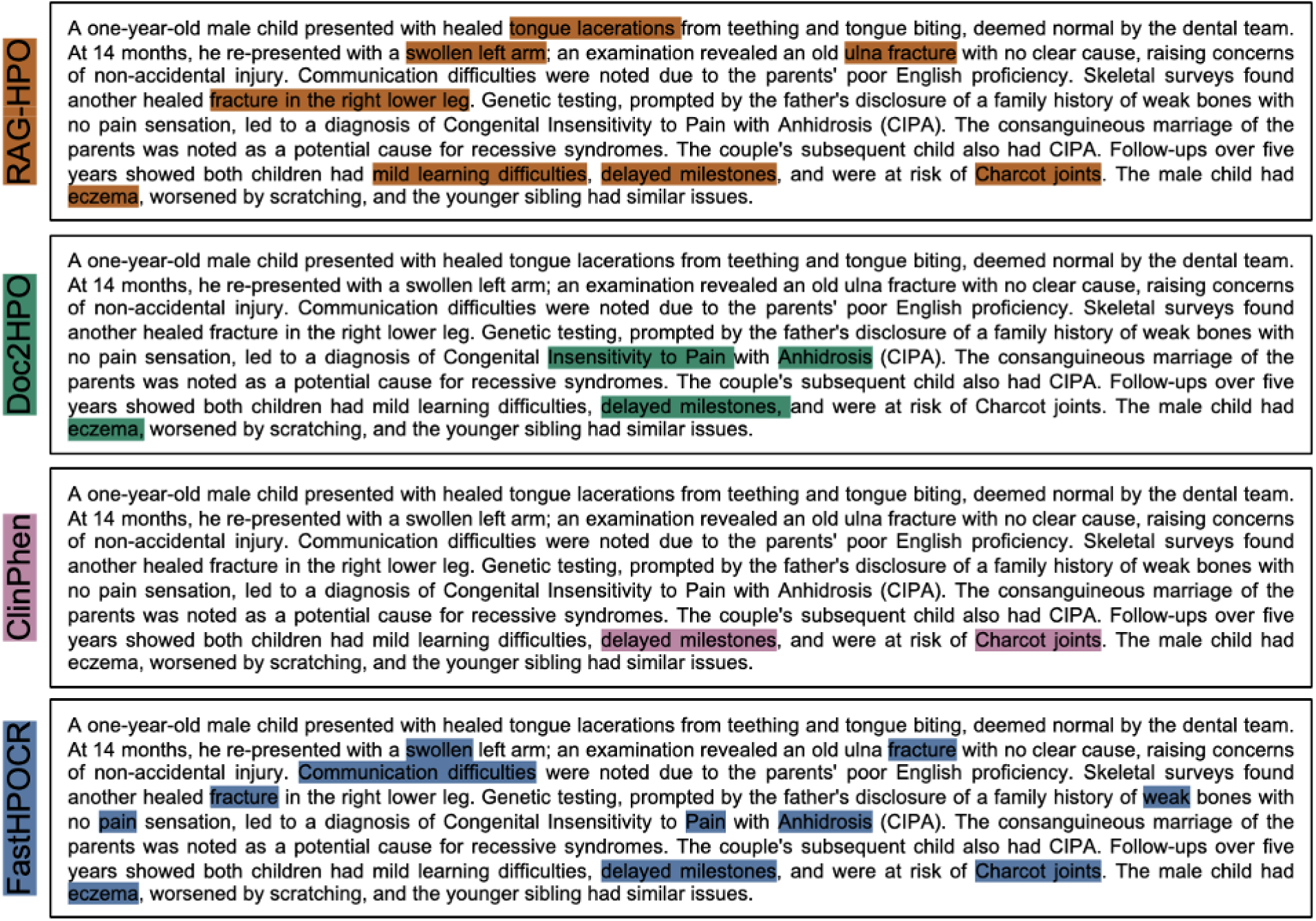
*Side-by-side comparison of HPO Analysis tools in a sample case.* The passage highlights show what phenotypic phrases are identified by each tool. The table demonstrates the HPO terms assigned by each tool to the passage along with their associated phrase. RAG-HPO identifies more phenotypes and assigns more phrases than Doc2HPO or ClinPhen, but fewer than FastHPOCR. However, the terms assigned by RAG-HPO are more precise and relevant than those assigned by FastHPOCR. The passage was adapted from Mughal *et al., Cureus* 2021. [29]

**Table 3.** RAG-HPO assigns relevant HPO terms with greater accuracy than other analysis tools.

| Identified Phrase | RAG-HPO | Doc2HPO | ClinPhen | FastHPOCR |
| --- | --- | --- | --- | --- |
| Healed Tongue Lacerations | *HP:0000206\** | -- | -- | -- |
| Swollen Left Arm | HP:0010742 | -- | -- | -- |
| Old Ulna Fracture | HP:0003987 | -- | -- | HP:0003987 |
| Healed Fracture Right Lower Leg | HP:0041081 | -- | -- | -- |
| Eczema | HP:0000964 | HP:0000964 | -- | HP:0000964 |
| Mild Learning difficulties | HP:0001328 | -- | -- | -- |
| Delayed milestones | HP:0001263 | HP:0001263 | HP:0001263 | HP:0001263 |
| Risk of Charcot Joints | HP:0002821 | -- | HP:0002821 | HP:0002821 |
| Anhidrosis | -- | *HP:0000970\** | -- | *HP:0000970\** |
| Pain | -- | *HP:0012531\** | -- | *HP:0012531\** |
| Swollen | -- | -- | -- | *HP:0000969\** |
| Recurrent Fracture | -- | -- | -- | *HP:0002757\** |
| Communication Difficulties | -- | -- | -- | *HP:0034434\** |
| Weak | -- | -- | -- | *HP:0025406\** |

In this example, RAG-HPO with Llama-3 70B correctly assigned 7 out of 8 HPO terms. In comparison, Doc2HPO correctly assigned 2 terms but falsely identified 2 others, ClinPhen correctly assigned 2 terms, and FastHPOCR correctly assigned 4 terms while falsely identifying 6 terms. False identifications are indicated by italicized font.

### Evaluation of False Positive Results Among HPO Analysis Tools

To better understand the meaning of these incorrectly attributed terms, we conducted a deeper analysis of the relationship of false positives to the manually annotated HPO terms, specifically focusing on the ontological connections between the HPO terms selected by the programs and those identified through manual annotation.

We categorized the false positive HPO terms into three distinct groups: (1) terms that were direct ancestors of the target HPO terms, (2) terms that were indirectly related to the target terms, and (3) terms that were completely irrelevant to the correct terms. Additionally, for RAG-HPO, we classified any false positive HPO terms that did not exist in the HPO database as “hallucinations” generated by the system (**Figure 5A).**

**Figure 5:**
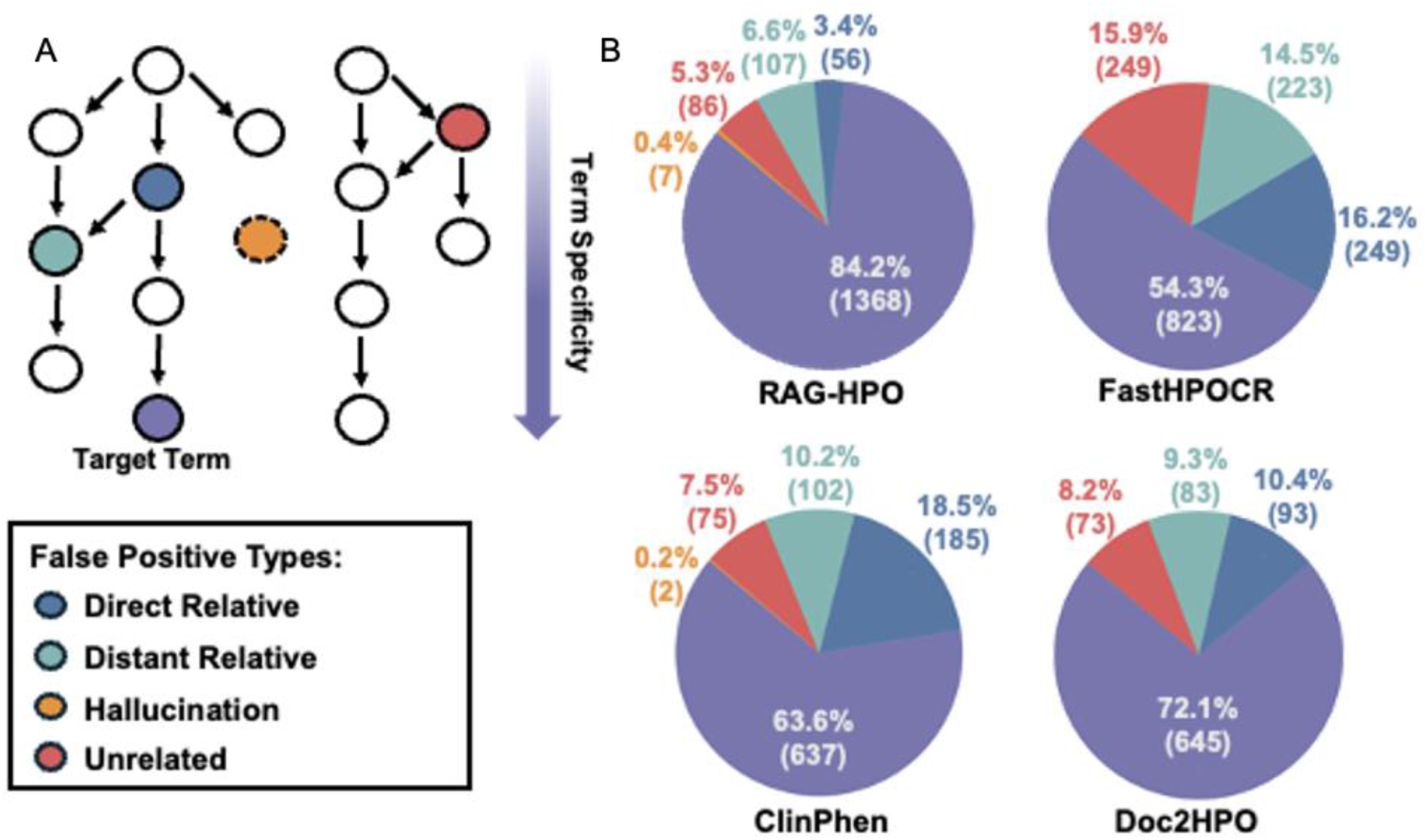
*Analysis of the relationship of assigned HPO Terms to manual standard.* Examination of the HPO terms returned that did not match the manually annotated standard were considered false positives. A) We completed a deeper analysis of these terms to better understand their nature and categorized them into distinct groups based on their relationship to the standard: Direct relatives, distant relatives, hallucinations, and unrelated terms. B) Depicts the false positive types in relation to the target term (in purple) with direct and indirect relatives being on the same phenotypic abnormality branch, unrelated terms belonging to completely different branches of the HPO, and hallucinations having no existence within the HPO database.

In our analysis, we accepted only the most specific term to describe each phenotype. However, there is value in identifying directly related terms, which are often broader descriptors of the phenotypes observed (**Figure 5A**). For example, a patient described as having a “swollen left arm” (HP:0010742) was assigned the term “swollen” (HP:0000969) by FastHPOCR—a directly related but less specific term (**Figure 4, Table 3**).

An example of indirectly related HPO terms is also shown in **Figure 4**, in which RAG-HPO identified “healed tongue lacerations” as a relevant phenotype but assigned the term HP:0000206 (glossitis), which refers to tongue inflammation. A better fit for this phenotype would have been HP:0034417 (intraoral laceration). These two terms are connected in the ontology through the broader term HP:0000163 (Abnormal oral cavity morphology).

In some instances, the tools assigned HPO terms that were unrelated to or not included within the manually annotated standard. For FastHPOCR, Doc2HPO, and ClinPhen, these errors often arose from terms mentioned in the case report that were not applicable to the specific patient. Such phrases typically included negated terms, components of syndrome names, or family history references that were not directly attributed to the patient. An example of this can be seen in **Figure 4**, where Doc2HPO and FastHPOCR incorrectly identified “Pain” (HP:0012531) and “Anhidrosis” (HP:0000970) as phenotypes for the patient, when these terms were part of the disease name “Congenital Insensitivity to Pain and Anhidrosis,” but were not explicitly stated as phenotypes for the patient.

Unrelated HPO terms assigned by RAG-HPO rarely included errors attributable to poor semantic similarity search. When RAG-HPO identifies a phenotypic phrase for term assignment, the phrase is vectorized and compared to the custom vector database we created. In rare cases, the search returned database entries that were completely unrelated to the extracted clinical phrases. As currently written, the LLM must choose an HPO term from the list generated by similarity search, a constraint designed to prevent hallucinations, but which can lead to nonsensical HPO choices.

Across the entire cohort, RAG-HPO hallucinated only 7 HPO terms, accounting for less than 3% of its false positives and an even smaller fraction of the total number of HPO terms assigned within the test cohort (**Figure 5B**). FastHPOCR and Doc2HPO produced no hallucinations, as expected due to their design. ClinPhen hallucinated 2 terms, likely due to obsolete HPO terms that had been retired from the database between the tool’s publication and our manual annotation of the cohort.

A deeper understanding of the false positive population helps us to better understand the reasoning for their development and determine ways to reduce their occurrence and improve our precision. This analysis also highlights the importance of understanding the level of detail needed for specific downstream applications. In some instances, less specific but related terms may be acceptable.

## Discussion

### Summary

We have developed RAG-HPO, a powerful tool for precise and comprehensive extraction of HPO terms from clinical case data using LLMs and RAG. RAG-HPO demonstrates superior precision and recall compared to existing tools for HPO analysis, offering greater flexibility and ease of use.

### Strengths

RAG-HPO stands apart from other LLM-based HPO analysis tools due to its flexibility and user-friendly design. It is accessible via a browser-based interface, similar to Doc2HPO, and can also be integrated into custom pipelines for broader analysis of patient data. This adaptability allows it to handle free-form medical text, making it especially useful in rare disease research, where patient descriptions often vary in format. RAG-HPO enables users to submit plain text without needing to preprocess data or integrate into electronic health records (EHR).

One of RAG-HPO’s key strengths is its compatibility with a variety of LLMs. For HIPAA compliance, users can pair it with locally stored LLMs using widely available software solutions. The primary requirement is that the LLM supports the OpenAI API framework, which ensures broad compatibility for sending and receiving queries and results.

A major advantage of RAG-HPO over traditional fine-tuning of LLMs is its computational efficiency and flexibility. Fine-tuning requires significant resources—both in terms of time and computational power—to retrain models, along with specialized expertise in generative AI. In contrast, RAG allows users to update the vector database quickly with minimal technical knowledge, requiring only a few clicks to ensure up-to-date results. This makes RAG-HPO far more accessible and practical for clinicians and researchers who may not have the capacity for extensive model fine-tuning.

Recently, others have used a pre-determined dataset called the HPO Gold Standardized Corpora (GSC) to evaluate their analysis programs. [13, 21] The GSC dataset is developed from entries to the Online Mendelian Inheritance in Man (OMIM) database and contains a wide variety of entries describing genetic diseases. [28] Though we also had access to this database, we ultimately chose to evaluate RAG-HPO on previously published case studies because the format of these case reports more closely resembled real clinical notes. Entries within the GSC dataset include some passages formatted like a clinical note, but also contained descriptive entries meant to educate individuals about a particular disease. Additionally, the manually annotated HPO lists (“truth sets”) of the GSC often assigned lower information-content HPO terms to phrases that were consistent with a more specific term, representing a loss of more detailed phenotypic information. This is likely due to the nature of the HPO database, which is consistently updated with terms and IDs that are more accurate for a given phenotype. Given these obstacles, we developed our own database of clinical case reports to test RAG-HPO and compared its performance with other programs as described. As we have demonstrated, RAG-HPO performs better at automated deep phenotype analysis compared to competitors.

### Limitations

One limitation of RAG-HPO is its processing speed. While individual clinical notes may take up to 45 seconds to analyze, other tools, such as FastHPOCR, can process the entire 112 case cohort in the time it takes RAG-HPO to do a single case. However, the longer analysis time for RAG-HPO is offset by its superior precision and comprehensive output, which significantly reduces the overall time required for accurate deep phenotyping of a patient case.

Another challenge is that the precision of RAG-HPO depends heavily on the quality of the vector database used for semantic similarity searches. Our vector database is meticulously maintained, with additions being made frequently to improve its capabilities. However, it is also limited by the available HPO IDs, which are tailored towards rare disease traits. As the Human Phenotype Ontology expands and the database is refined, precision is expected to improve. However, the subjective nature of phenotype interpretation can still influence results, as different users may prioritize varying levels of specificity. While RAG-HPO is designed to identify the most precise HPO term for a given phenotypic phrase, some users may seek a broader characterization of a patient’s phenotype.

The inherent subjectivity of clinical evaluations, through which different physicians may arrive at slightly different interpretations of the same patient, can also impact the phenotyping process. Although HPO terms aim to standardize patient presentations, variability in clinical assessment may still affect the final set of phenotypes attributed to an individual patient. While minimized with the use of LLMs in the automated deep phenotyping process, this subjectivity will persist when comparing cases of individuals seen by different physicians. Further downstream analyses focused on comparing HPO term lineages along with the specific HPO terms can further mitigate the impact of clinician subjectivity in patient evaluation.

RAG-HPO’s recall is contingent on the LLM used in conjunction with the program. Higher-powered LLMs trained with more information offer greater accuracy in identifying relevant phenotypes and assigning HPO terms because of their greater ability to understand and interpret natural language and context. Fortunately, RAG-HPO is built to allow users flexibility in their choice of model. We have selected Llama-3 70B, a mid-range open-source LLM, as a suitable option that balances performance with accessibility. Given the rapid pace of advancements in generative AI, we anticipate continued improvements in model performance.

### Future Directions

Our primary objective in developing RAG-HPO is to create an efficient and user-friendly tool for automated deep phenotyping. Looking ahead, we plan to further improve the vector database by incorporating at-large user contributions of verified phrases that consistently match establish HPO IDs. While RAG-HPO was initially designed for deep phenotyping in rare variant discovery, its potential applications could extend to include other applications within the medical field that require the comparison or identification of patients based on their clinical presentation. Future efforts will focus on expanding these applications to broaden the utility of RAG-HPO in clinical and research settings.

## Conclusions

RAG-HPO represents a significant advancement in the field of automated deep phenotyping, demonstrating a marked improvement over traditional HPO tools in both precision and recall. By leveraging Retrieval-Augmented Generation (RAG) and a dynamic vector database of over 54,000 phenotypic terms, RAG-HPO offers clinicians and researchers an accessible, computationally efficient solution for phenotype extraction that is both adaptable and resource-light. Unlike conventional LLM-based tools, RAG-HPO minimizes the need for fine-tuning and reduces the risks of “hallucinations,” making it a reliable and scalable tool for phenotypic analysis, particularly in rare disease diagnosis and clinical genomics.

## Data Availability

All data produced are available online at https://github.com/PoseyPod/RAG-HPO

## List of abbreviations

API: Application Programming Interface
GSC: gold standardized corpora
HPO: human phenotype ontology
JSON: JavaScript Object Notation
LLM: large language model
OMIM: Online Mendelian Inheritance in Man
RAG: retrieval augmented generation

## Additional Files

### Additional File 1

CSV file

Vector Database Dictionary

This spreadsheet file contains a table of all key-value pairs, mapping each clinical word or phrase (‘key’) to an HPO ID (‘value’).

### Additional File 2

DOC file

Supplemental Text and Table

This text file includes supplemental text demonstrated the system messages generated by RAG-HPO and provided to the user, as well as Table S1.

### Additional File 3

XLS file

RAG-HPO Tests and Data Analysis

This spreadsheet file contains the HPO IDs for the case sets against which benchmarking was performed, as well as the benchmarking data itself, across all tested tools.

## Declarations

### Ethics approval and consent to participate

all clinical texts and data used herein were derived from published case reports available in the medical literature or the published gold standardized corpora. No unpublished clinical or research patient data were implemented in this study.

### Consent for publication

Not applicable. No unpublished clinical or research patient data were implemented in this study.

### Availability of data and materials

Published case reports and data used to benchmark RAG-HPO are provided within the Supplemental Files. RAG-HPO is available for download at https://github.com/PoseyPod/RAG-HPO

### Authors’ contributions

BTG designed the study, created the software, collected, analyzed and interpreted the data, and drafted the manuscript. PY selected the test cohort independently of other authors. BTG and LW created the manually annotated standard. LW, PY, NG, EARM, HD, MD, and AJ assisted with data collection, analysis, and interpretation. JRL assisted with data interpretation. JEP supervised the study design, data collection and analysis. All authors revised the manuscript and approved the final submission.

## Acknowledgements

We would like to thank the following for their help and support in creating RAG-HPO: Thanks to Steve Ludke, PhD, Maxim Seferovic, PhD, and Shinya Yamamoto DVM, PhD for your guidance in coding in Python; thanks to Gwendolyn Hummel for biostatistics guidance; and thanks to Cole Deisseroth for being a resource for information on HPO analysis and the application of NLP to deep phenotyping.

